# A Machine Learning Model Integrated with the Clinical Workflow Detects Sepsis Early with High Sensitivity and Specificity

**DOI:** 10.1101/2023.08.17.23294209

**Authors:** Mohammed A. Mahyoub, Ravi R. Yadav, Kacie Doughetry, Ajit Shukla

**Affiliations:** Virtua Health, Marlton, New Jersey 08053; Binghamton University, Binghamton, New York, 13902

**Keywords:** Sepsis, Early detection, Machine learning, XGBoost, Model interpretability, Machine learning deployment

## Abstract

**Background:** Sepsis is a life-threatening condition caused by a dysregulated response to infection, affecting millions of people worldwide. Early diagnosis and treatment are critical for managing sepsis and reducing morbidity and mortality rates.

**Materials and Methods:** A systematic design approach was employed to build a model that predicts sepsis, incorporating clinical feedback to identify relevant data elements. XGBoost was utilized for prediction, and interpretability was achieved through the application of Shapely values. The model was successfully deployed within a widely used Electronic Medical Record (EMR) system.

**Results:** The developed model demonstrated robust performance pre-operations, with a sensitivity of 92%, specificity of 93%, and a false positive rate of 7%. Following deployment, the model maintained comparable performance, with a sensitivity of 91% and specificity of 94%. Notably, the post-deployment false positive rate of 6% represents a substantial reduction compared to the currently deployed commercial model in the same health system, which exhibits a false positive rate of 30%.

**Conclusions:** These findings underscore the effectiveness and potential value of the developed model in improving timely sepsis detection and reducing unnecessary alerts in clinical practice. Further investigations should focus on its long-term generalizability and impact on patient outcomes.

## 1. Introduction

Sepsis is a life-threatening organ dysfunction resulting from a dysregulated host response to infection that impacts millions of people around the world [1]–[3]. Early diagnosis and treatment are crucial in managing sepsis, a life-threatening medical condition leading to increased morbidity and mortality rates [4]. There are some established scoring systems for assessing the risk of sepsis [5]–[8]. These systems assign risk scores based on a set of predefined criteria. Due to the complexity of sepsis and its nonlinear association with patient characteristics (e.g., vitals and labs), these tools are limited in terms of distilling predictive insights from the patient profile. Therefore, there has been a growing interest in using machine learning algorithms as decision-support tools for sepsis detection [9]–[12]. While there is an abundance of articles on the topic, there is still a need for improving the interpretability of models and investigating model deployment in operation in hospitals.

Machine learning as a concept was introduced in 1959 by Arthur Samuel in the quest to grant computers the ability to learn without being explicitly programmed [13]. It has been proven to be very beneficial in many medical and healthcare delivery applications including the early detection of diseases, robotic-assisted surgeries, extracting insights from clinical text (e.g., reports, physician notes, etc.), and labeling medical images with potential diagnosis [14]–[20]. Predictive modeling in particular can serve as a decision-support tool in clinical settings [21]– [25]. Generating predictive scores for patients can lead to better prioritization of healthcare delivery, early appropriate interventions, and streamlining patient care workflows in general.

Machine learning models have been increasingly considered for predictive modeling research in healthcare. However, most published articles have not reported successful implementations of proposed models, leading to a huge gap between theory and practice. This fact emphasizes the inevitable need for practical research that addresses the predictive model lifecycle from conceptualization to operationalization. In this study, we present an end-to-end machine learning model for the early detection of sepsis, enlightening the adoption and integration of the proposed model within the clinical workflow.

This study presents an operationalizable machine-learning model that consolidates clinical data from different sources (e.g., vitals, labs, medications, etc.) to detect sepsis early, allowing for early interventions and administration of the health of patients. In this paper, we make four main contributions:

- It proposes a framework for identifying, mapping to the database, and examining with clinical subject matter experts a list of sepsis predictive data elements.
- It proposes an operationalizable machine-learning model for the early detection of sepsis with high sensitivity and specificity.
- It integrated the prediction model with the clinical workflow through the integration of the model with a widely adopted EMR system in the U.S.
- It provides an interpretability and explainability analysis of the machine learning model.

## 2. Materials and Methods

### 2.1. Research Methodology Overview

We adopted a systematic approach for conducting the research and building the models in this paper. The institutional review board (IRB) of Virtua Health approved this study. Virtua Health IRB waived the requirement for consent because the research involved no more than minimal risk to subjects, could not practically be carried out without the waiver, and the waiver will not adversely affect the rights and welfare of the subjects. This requirement of consent was waived on the condition that, when appropriate, the subjects will be provided with additional pertinent information about participation. Figure 1 illustrates the overall process. We began by evaluating the performance of a commercially available sepsis prediction model that was integrated in Virtua Health’s EMR system. We identified issues present in this commercially available model, and used that as a launching point for developing a new sepsis prediction model. After pinpointing these problems, we conducted a comprehensive analysis of sepsis prediction models in the literature and extensive discussions with the clinical team. Based on these investigations, we identified a list of potential predictive data elements for sepsis, validated the list with clinical subject matter experts, mapped the data elements to the clinical database, and developed SQL queries to retrieve the data from the various sources (i.e., tables) in the database. Then, we cleaned, preprocessed, transformed, and performed feature engineering on the retrieved dataset. Based on the resulting dataset, we developed and trained the machine learning model. The model was evaluated through error and classification performance analyses. After that, we prepared the model artifacts for cloud deployment and integration with the EMR system. Then, we collected post-deployment performance data to evaluate the deployed version of the model. Finally, we set up the monitoring pipeline for the model in operation.

**Figure 1.**
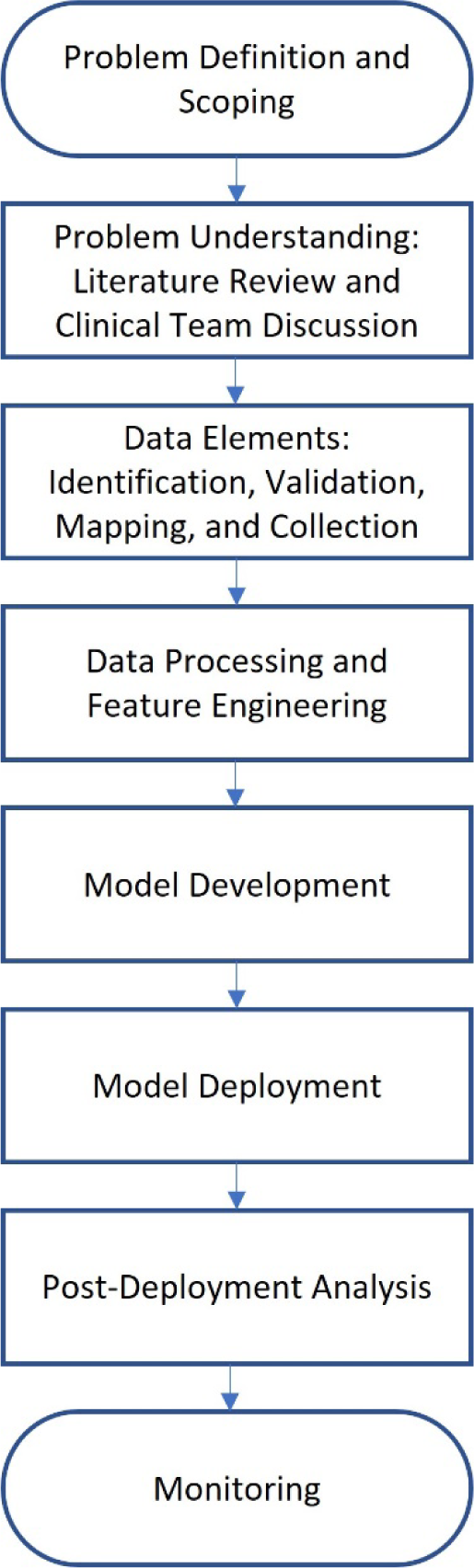
Research methodology workflow.

### 2.2. Data Identification and Collection

To construct the sepsis prediction model, we conducted a literature review to determine potential data elements [10], [26], [27]. We compiled an initial list of data elements from various categories such as demographics, vitals, labs, medications, and lines, drains and airways (LDAs). Then, we consulted with clinical subject matter experts to gain insight into the data and solicit feedback and additional data elements from their perspectives. We performed a mapping analysis on the final list of data elements, in which we examined the correspondence of the data elements to the data items in the EMR’s front end (i.e., Epic hyperspace). We also generated a related list of tables and columns in the backend database from these mappings. This analysis is essential for obtaining the appropriate data elements for predictive modeling. Next, we used advanced SQL queries to extract the data from different sources in the Clarity database of the Epic EMR. We collected data for patients who were 18 years or older and who were admitted as inpatients at one of Virtua Health’s five hospitals (Virtua Marlton Hospital, Virtua Mount Holly Hospital, Virtua Our Lady of Lourdes Hospital, Virtua Voorhees Hospital, and/or Virtua Willingboro Hospital). We restricted the patient population to those who were admitted to the inpatient hospitals between January 2020 and July 2022. The initial cohort for model training and validation consisted of 17,750 inpatient encounters (70% for training the model and 30% for validation). See Section 3.1 for descriptive statistics of the dataset. The model’s post-deployment performance was evaluated by collecting model scores and patient final DRGs from the Clarity database. The sepsis label was assigned to patients based on their DRGs. Patients with DRGs 870, 871, or 872 were considered septic. DRG 870 is for Septicemia or Severe Sepsis with Mechanical Ventilation (MV) >96 Hours. DRG 871 is for Septicemia or Severe Sepsis without MV >96 hours and with Multiple Chronic Conditions (MCC). DRG 872 is for Septicemia or Severe Sepsis without MV >96 hours and without MCC.

### 2.3. Variable Selection and Data Processing

Data processing was performed on the retrieved dataset to prepare it for machine learning modeling. We adopted several steps of analysis and processing including data encoding, imputation, aggregation, and scaling.

The original dataset contained around 86 variables spanning several categories such as demographics, vitals, initial sepsis screening, labs, medications, LDAs, and comorbidities. Demographic variables are Age, Gender, Is_Married, Emergency Department (ED) Length of Stay, and Inpatient Length of Stay. Vitals variables are Temperature, Respiratory Rate, Systolic Blood Pressure, Diastolic Blood Pressure, Mean Arterial Pressure, Heart Rate, and Oxygen Saturation. Initial sepsis screening in the ED variables are Level of Consciousness, Are there Two or More Signs of Sepsis, and History of Sepsis. LDA variables are the Count of Central Venous Catheters, Count of Closed Suction Drains, Count of Endotracheal Tubes, Count of Feeding Tubes, Count of Incisions, Count of Peripheral IVs, Count of PICCs, Count of PORTs, Count of Pressure Ulcers, Count of SWAN-GANZ Catheters. Labs are Absolute Lymphocyte Manual Count, Absolute Neutrophil Manual Count, Base Excess Arterial, Calcium, Chloride, CO2, Creatinine, FiO2, Glucose, HCO3, Hematocrit Automated Count, Hemoglobin, Hemoglobin A1C, Lactate, Lymphocyte Automated Count, Magnesium, Mean Corpuscular Hemoglobin Concentration, Monocyte Automated Count, Neutrophil Automated Count, Nucleated RBC Automated Count, PaCO2, pH Blood, Phosphate, Platelet Automated Count, POC Glucose, Potassium, Procalcitonin, RBC Automated Count, RBC Distribution Width Automated Count, RBC Morphology, Reticulocyte, and WBC Automated Count. Counts of the following medications: Alpha Beta Blockers, Analgesic Antipyretics, Antianginals, Antifungals, Antihypertensives, Beta-Adrenergic Agents, Beta Blockers, Beta Blockers Cardiac Selective, Betalactam Antibiotics, Cephalosporins, Coronary Vasodilators, Fluoroquinolones, Glucocorticoids, Leukocyte Stimulators, Narcotic Analgesics, Penicillins, Proton Pump Inhibitors, Sodium Saline, and Vancomycin and Glycopeptides. Comorbidities are Chronic Kidney Disease, Chronic Liver Disease, Congestive Heart Failure, COPD, Coronary Artery Disease, Diabetes, HIV, Hypertension, and Obesity.

Some variables in the dataset are categorical. For example, the Is_Married variable can take two possible values: Yes or No, and the Gender variable can have values such as Male and Female. Therefore, we performed data encoding to transform the categorical values into numerical values for machine learning modeling. This step is because all machine learning algorithms operate on numerical data and computations. In this study, we applied label encoding, which is a common technique in machine learning literature. This technique assigns a distinct numerical value to each value of a categorical variable. A specific example of encoding the Is_Married variable is to assign 1 to the value “Yes” and 0 to the value “No”.

Many variables in the data set were collected hourly, and the data collection resulted in data sparsity. To address this challenge, we calculated the mean value of each variable. For example, for a given encounter, we aggregated the features’ data before the sepsis onset or recording time, resulting in a one average value per feature. The purpose of this dataset was to develop a sepsis recognition model. We hypothesized that a model that can recognize septic characteristics hidden in the large dataset, obtained by the aggregation process, can help identify sepsis before onset if the model is applied to patient profile regularly. The model may detect that a patient’s profile at a certain time during the stay resembles a septic profile and will generate an alarm. As shown in the Results section, the recognition model was evaluated on its ability to identify sepsis at a specific number of hours before onset. A simulation analysis was conducted to determine this performance. In summary, the sepsis model was trained on the aggregated dataset that was further processed to handle null values and the scaling issue as discussed below.

As stated previously, the original dataset has many null values due to the hourly collection schema that was used, resulting in sparsity. Even after aggregating the dataset using averages, some variables still have a large proportion of missing values. Therefore, we decided to eliminate only the extremely sparse variables. We experimented with different thresholds and settled on 90%. This means that any variable that has 90% or more missing values is removed from the dataset. This threshold might seem high, but we reasoned that for some patients, a variable that is mostly null for the whole population might be very valuable in detecting sepsis if it has a value for those specific patients. In other words, we allow the model to learn from the patterns of missing values in the dataset. The machine learning algorithm that we chose for the sepsis prediction model can handle datasets with null values well.

To avoid the bias and inefficiency caused by the heterogeneity of scales among the variables in the aggregated dataset, we applied a MinMax transformation to standardize all variables to a common range of (1, 5) as shown in Equation 1.

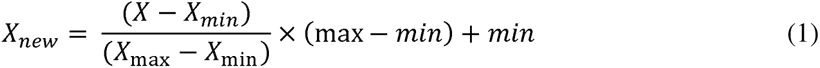

where *X_new_* denotes the standardized variable, *X* denotes the original variable, *X_min_* and *X_max_* denote the minimum and the maximum values of the original variable respectively, *max* and *min* denote the upper and lower bounds of the standardized range (i.e., 5 and 1 respectively).

### 2.4. Model Development

We used predictive modeling [28] to assign sepsis risk scores to patients based on their features. For sepsis prediction, the sepsis score and the patient’s characteristics have a complex and nonlinear relationship, which necessitates the application of an advanced machine learning algorithm. In this paper, we employed the Extreme Gradient Boosting algorithm (XGBoost).

XGBoost is a state-of-the-art algorithm for predictive modeling that employs the tree-boosting framework [29]. Tree boosting is a machine learning technique that combines multiple weak learners into a strong learner. The weak learners are usually decision trees that have a limited depth or number of leaves. The idea is to train the trees sequentially, each one trying to correct the errors of the previous ones. The final prediction is obtained by a weighted vote of the individual trees. In this paper, we used the XGBoost package in Python, which is a scalable and efficient implementation of tree boosting. XGBoost has several advantages over other boosting methods, such as regularization, parallelization, and handling of missing values. We tuned the hyperparameters of XGBoost using grid search. Grid search is a method for finding optimal or near-optimal combinations of hyperparameters for machine learning algorithms. Hyperparameters are parameters that set the configuration of the learning process and cannot be updated or adjusted using the training data itself. The hyperparameters that were changed, through hyperparameter tuning, from default values in the XGBoost package in Python are presented below in a key-value format: { “colsample_bytree”: 0.1, “gamma”: 4, “learning_rate”: 0.08, “max_delta_step”: 5.24, “max_depth”: 0, “min_child_weight”: 97, “n_estimators”: 255, “reg_alpha”: 18, “reg_lambda”: 2, “sampling_method”: “gradient_based”, “tree_method”: “hist”}.

### 2.5. Explainability Analysis

To interpret the predictions of our machine learning model, we applied SHAP (SHapley Additive exPlanations), a framework that computes feature importance values for each prediction [30]. SHAP values are derived from Shapley values, which are a fair allocation of the payoffs among the players in a cooperative game. In our context, the features were the players and the prediction outcomes were the payoffs. SHAP allowed us to measure how each feature influences the prediction, in a positive or negative direction, and also capture the feature interactions.

### 2.6. Model Deployment

We implemented the model into the clinical workflow to enhance its usability by clinicians. For this purpose, we hosted the model on Epic Nebula (Epic Systems, Verona, WI USA) and integrated it with the EMR system (Epic Systems, Verona, WI USA) (see Figure 2). The model artifacts were configured and tested using the Epic Slate Environment (Epic Systems, Verona, WI USA). This integration allowed for seamless data transfer from the Chronicles database to the model in the cloud for sepsis scoring and score delivery back to the system for alert generation and dashboard visualization. Input data was provided to the model through workbench reports that were customized for the sepsis model. We configured a batch job to execute the model every hour.

**Figure 2.**
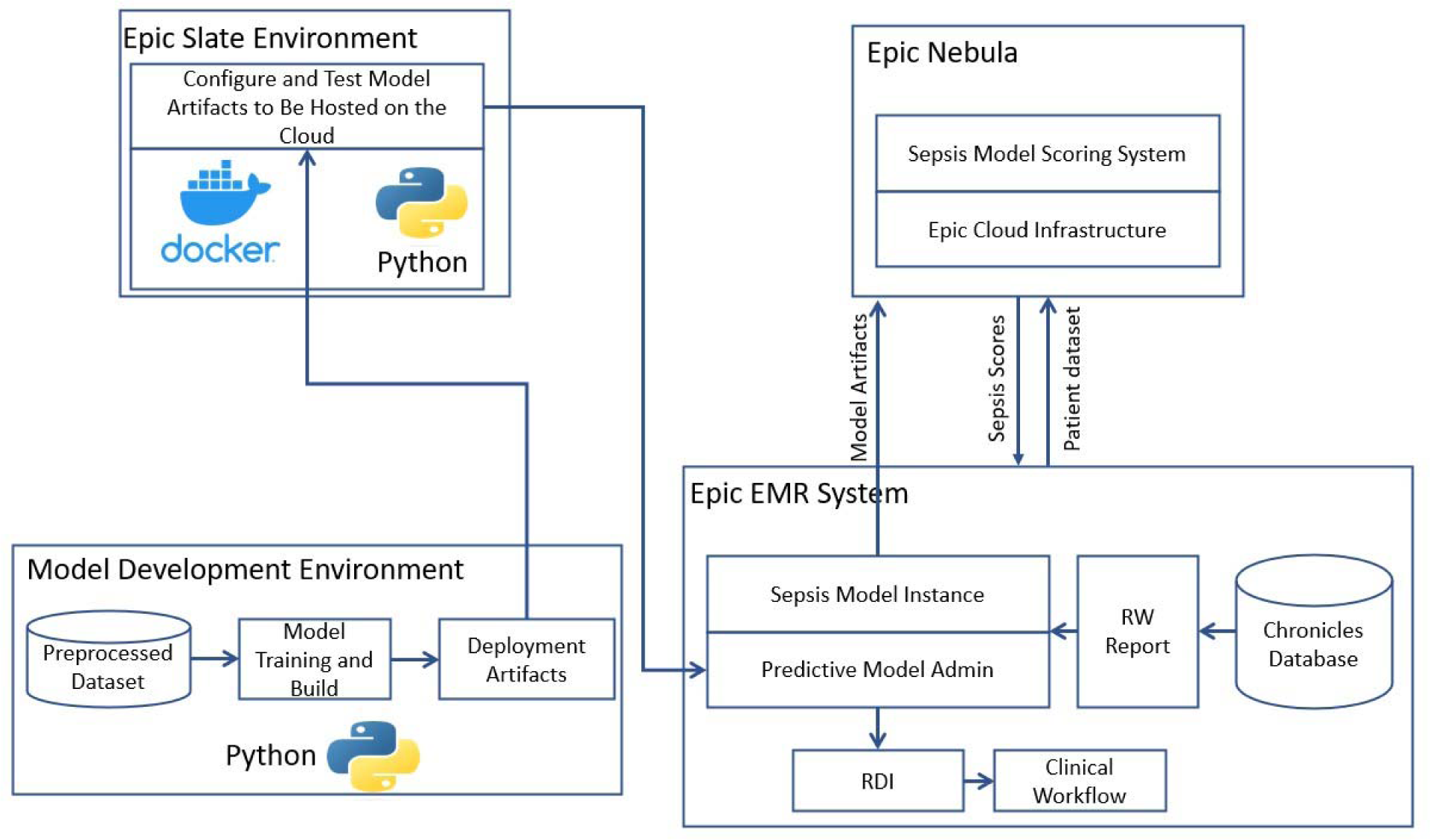
Model deployment and operationalization.

## 3. Results and Discussion

In this study, we built a model to predict sepsis in the hospital by training a model on retrospective EMR data, consisting of elements related to demographics, labs, vitals, etc. Importantly, each case was known to either include a sepsis diagnosis or did not. Table 1 presents the fundamental characteristics of the population. The population in the sepsis prediction model training dataset consisted of f 17,750 observations, representing a diverse group of individuals. The average age in the dataset was 54.83 years, with a standard deviation of 25.27 years, indicating a wide range of age groups. In terms of sex, the dataset shows a relatively equal distribution, with males accounting for 44% and females for 56% of the population. About 34% of the individuals in the dataset were married. The dataset also included information on hospital stays, with an average inpatient length of stay (LOS) of 207.16 hours, accompanied by a high standard deviation of 988.76 hours, suggesting significant variation in this measure. Additionally, the dataset includes data on ED visits, with an average LOS of 4.26 hours and a standard deviation of 3.53 hours. These characteristics and other clinical data elements reflect the diverse demographics and healthcare experiences captured in the training dataset, providing valuable information for the machine learning algorithm to learn and make predictions.

**Table 1.** Demographics of the population in the training dataset.

| Characteristic | Value<br>mean (std) |
| --- | --- |
| Total number of observations | 17,750 |
| Age | 54.83 (25.27) years |
| Gender | Male: 44%; Female: 46% |
| Is Married? | 34% |
| Inpatient Length of Stay (LOS) | 207.16 (988.76) hours |
| Emergency Department LOS | 4.26 (3.53) hours |

We trained a sepsis prediction model using the XGBoost algorithm and used it to predict whether a case included a sepsis diagnosis in our held-out testing dataset. We then compared the model’s prediction of sepsis or no sepsis to the known sepsis or no sepsis diagnosis from the EMRs. Figure 3 displays the confusion matrix of the sepsis prediction model on the test dataset, generated using a sepsis score threshold of 0.05 (a score above 0.05 means sepsis and below means no sepsis). The counts of true positive (TP), true negative (TN), false positive (FP), and false negative (FN) predictions are presented in Figure 3. The sepsis prediction model demonstrated a sensitivity (true positive rate) of 92%, indicating 92% of the sepsis cases in the test dataset were correctly identified by the model. The sepsis prediction model demonstrated a specificity (true negative rate) of 93%, indicating 93% of the non-sepsis cases in the test dataset were correctly identified by the model. The false positive rate, calculated as 1 - specificity, was 7%. This signifies that the model incorrectly predicted sepsis in 7% of the cases that were truly negative for sepsis. While a lower false positive rate is desirable, the combination of high sensitivity and specificity suggests that the model performs well in accurately predicting sepsis and distinguishing it from non-sepsis cases. Overall, the high sensitivity and specificity achieved by the sepsis prediction model in Figure 4 contribute to its accurate identification of sepsis cases. A high sensitivity ensures that a significant number of true positive sepsis cases are correctly detected, while a high specificity guarantees a low number of false positive predictions. These attributes are crucial in enabling healthcare professionals to promptly identify sepsis cases and administer appropriate interventions efficiently, thereby enhancing patient care and outcomes.

**Figure 3.**
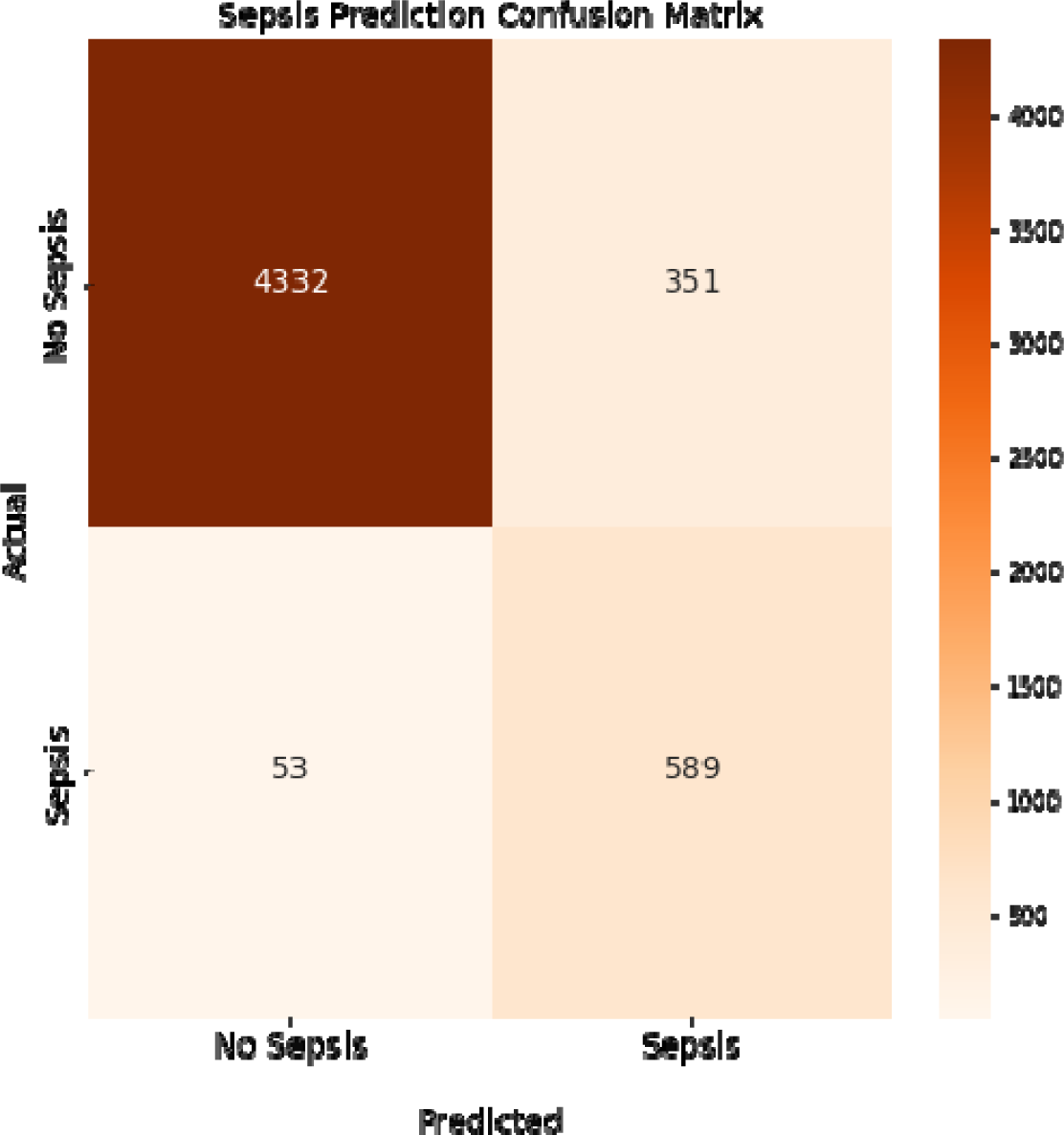
Confusion matrix of the sepsis prediction model. Sensitivity is 92% and specificity is 93% with a 7% false positive rate. Colorbar represents the count of cases.

**Figure 4.**
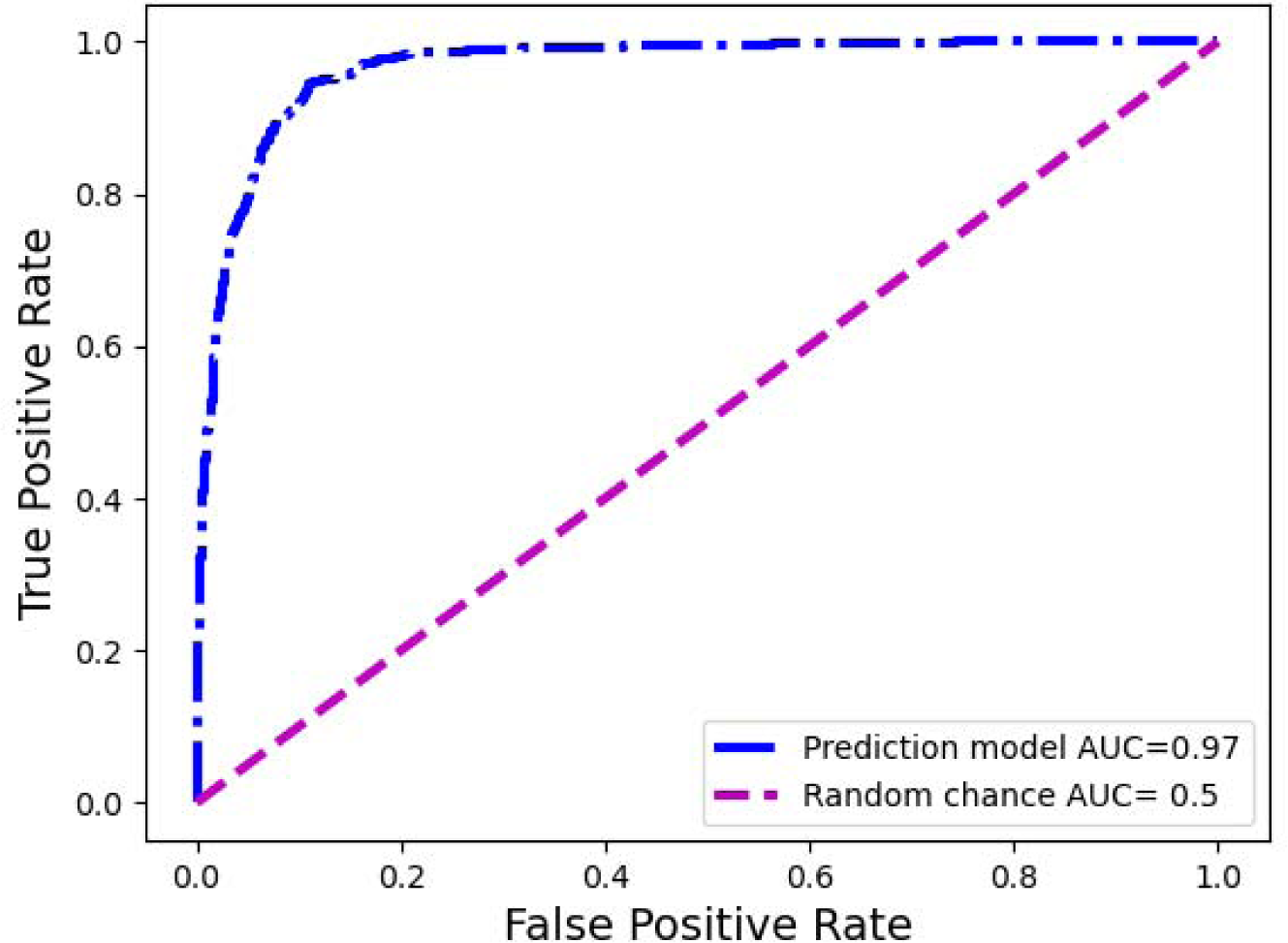
Receiver operating curve of the sepsis prediction model: Pre-deployment performance

The ROC curve is a graphical representation of the performance of a binary classification model, such as the sepsis prediction model in this context. It demonstrates the trade-off between the true positive rate (sensitivity) and the false positive rate (1-specificity) for various classification thresholds. In the case of sepsis prediction, the ROC curve showcases the model’s ability to correctly identify patients who have sepsis (true positives) while minimizing the number of false positives. Figure 4 presents the receiver operating characteristic (ROC) curve of the sepsis prediction model, showcasing its performance prior to implementation. The curve is derived from the testing dataset, which comprises 30% of the available data used for model development.

The area under the curve (AUC) for this model is an impressive 0.97, indicating a high level of accuracy and discrimination (correctly predicting cases of sepsis and correctly predicting cases with no sepsis) in predicting sepsis. With an AUC of 0.97, the sepsis prediction model demonstrated exceptional performance in distinguishing between sepsis and non-sepsis cases. The higher the AUC value, the better the model’s ability to differentiate between the two classes. This implies that the model exhibits a high sensitivity in identifying sepsis cases while maintaining a low false positive rate. Consequently, this model holds promising implications for accurately predicting sepsis, providing healthcare professionals with a valuable tool for early intervention and improved patient outcomes.

After developing a model with high sensitivity and specificity, we investigated which features (predictors) contribute most to the sepsis prediction score. We approached this by computing Shapley values. Figure 5 provides valuable insights into the Shapley values of features and their correlation with the sepsis score. The Shapley values represent the contribution of each feature to the final prediction. The feature range values are visually depicted through a color spectrum from blue to red, indicating low to high values, respectively. The grey color represents missing values (i.e., NaN). The x-axis represents the Shapley values, while the alignment of the feature violin plots illustrates how the feature values influence the sepsis risk score. Positive Shapley values indicate that corresponding feature values drive the sepsis scores towards 1, implying a greater likelihood of sepsis in patients with such values. Notably, Figure 5 highlights that Lactate is the most important predictor in this dataset, as higher Lactate values correspond to a higher risk of being septic. The figure goes on to list the top 20 most important features and their contribution to the sepsis score. By examining Figure 5, healthcare professionals and researchers can gain insights into the factors that significantly influence the prediction of sepsis. Understanding the impact of specific features on the sepsis risk score enables healthcare providers to identify critical indicators and prioritize appropriate interventions for patients who may be at a higher risk of developing sepsis. For example, it may be worth prioritizing acquiring lab results on lactate and neutrophil count in a timely manner so that the model can assess the risk of sepsis early.

**Figure 5.**
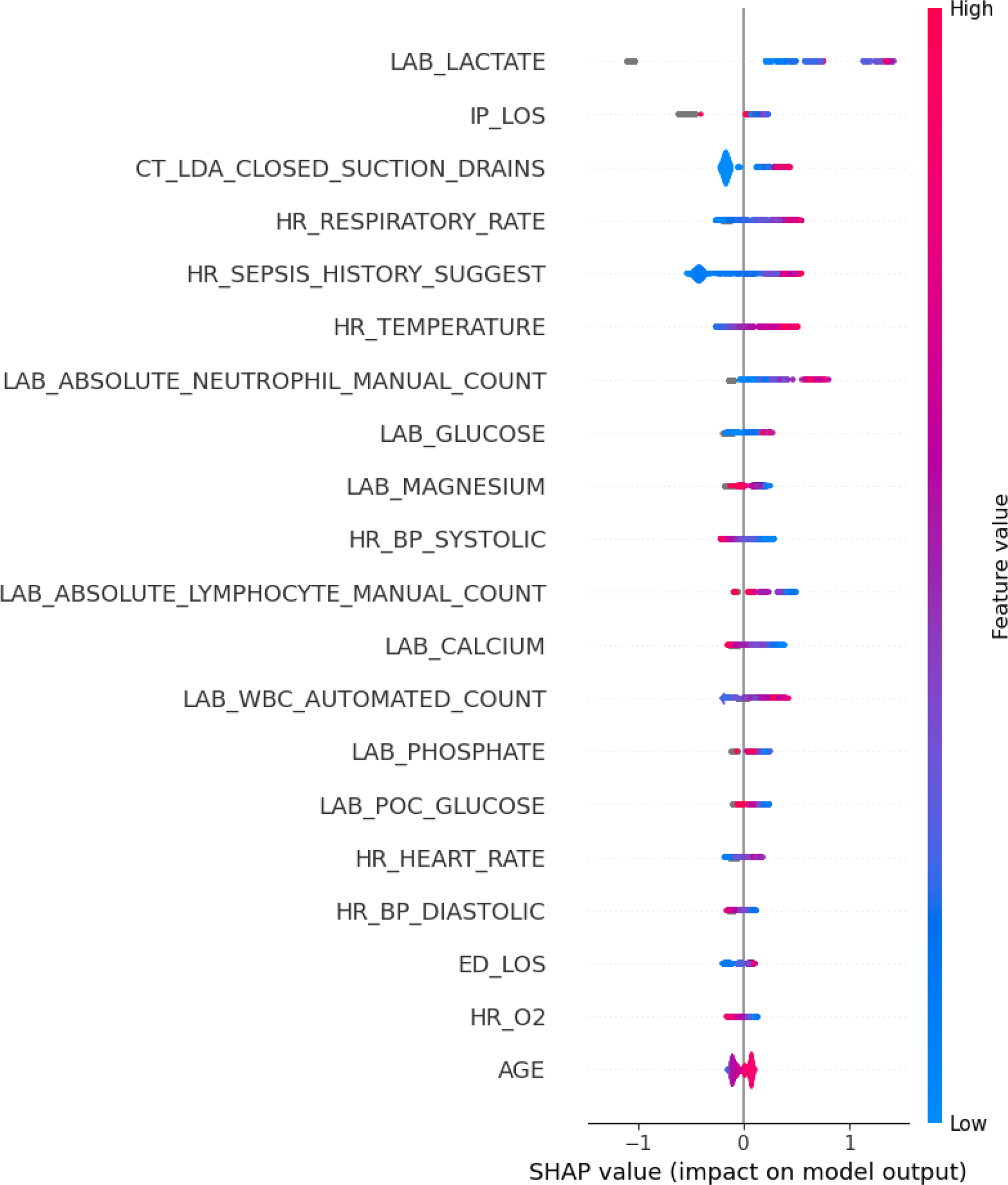
Highly important features and their corresponding Shapley values. The grey color depicts the missing values (NaN) in a certain feature.

A major aim of this study was to investigate the performance of a sepsis predictive model in operation in the hospital. Figure 6 illustrates the performance of the sepsis prediction model after deployment in operational settings. Post-deployment performance was based on a data collected from the system (Five hospitals) during operations (three-week period), comprising records of 1,896 patients, including 98 septic and 1,798 non-septic cases. It displays the sensitivity and specificity of the model at various thresholds ranging from 0.02 to 1, with increments of 0.01. The performance of the model in operations aligns closely with its pre-deployment performance, exhibiting a sensitivity of 91% and a specificity of 94% at a 0.05 threshold. These high values indicate the model’s ability to accurately identify both septic and non-septic patients. Moreover, the model achieves a low false positive rate of 6%, implying that only a small portion of non-septic patients are incorrectly flagged as septic. This low false positive rate has significant implications for patient outcomes, as it reduces the burden of nurse fatigue and minimizes the need for clinicians’ continuous checking due to false alarms. Comparatively, the false positive rate of 6% achieved by our sepsis model is substantially lower than that of a commercially available model adopted within the organization (30%). In other words, with our new sepsis predictive model, we reduced the number of false positive cases by 80%. This remarkable reduction demonstrates the superiority of our model in terms of minimizing unnecessary alerts and ensuring that healthcare providers can focus their attention on patients who genuinely require intervention for sepsis. The high performance of the sepsis prediction model, with its low false positive rate and high sensitivity, not only improves patient outcomes but also enhances the efficiency of healthcare professionals. Importantly, our model which was trained on a large retrospective dataset, maintains high specificity when running in the hospital setting.

**Figure 6.**
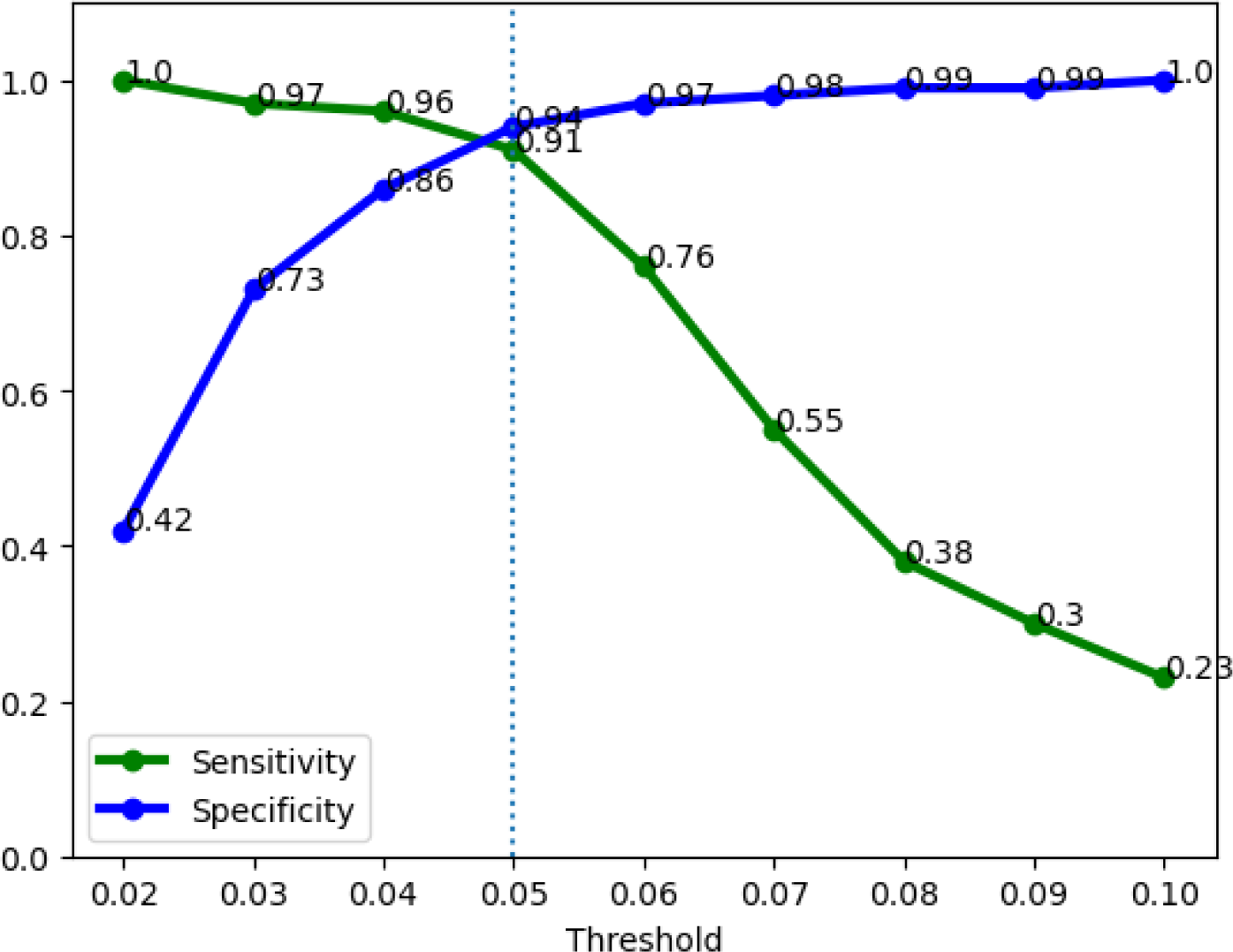
Operations performance of the sepsis prediction model. 91% sensitivity and 94% specificity at a 0.05 risk threshold.

## 4. Conclusions

In conclusion, the systematic design approach employed in this study, integrating clinical feedback and utilizing XGBoost with interpretability techniques, led to the successful development and deployment of a robust sepsis detection model. The model demonstrated high sensitivity and specificity pre-operations. Importantly, it maintained comparable performance after deployment, with a significantly reduced false positive rate compared to the currently deployed commercial model. These results highlight the potential value of the developed model in improving sepsis management and reducing alert fatigue in clinical practice.

Future research should address several important areas. Firstly, investigations should focus on assessing the generalizability of the model across different healthcare settings and patient populations, considering variations in clinical practices and data availability. Secondly, long-term studies are needed to evaluate the impact of the model on patient outcomes, such as mortality rates, length of hospital stay, and resource utilization. Additionally, efforts should be made to enhance the interpretability of the model further, enabling clinicians to better understand and trust its predictions. Moreover, further studies exploring the integration of the model with clinical decision support systems or real-time monitoring tools could provide valuable insights into its practical implementation and effectiveness.

## Data Availability

Data in the present study are not available due to agreements made with the IRB of Virtua Health.

